# Longitudinal immune transcriptomic signatures are associated with carotid intima–media thickness over 18 years

**DOI:** 10.64898/2026.03.12.26348144

**Authors:** Anna D’Antuono, Giacomo Fantoni, Elisabetta Giordano, Valentina Spigoni, Federica Fantuzzi, Francesca Bagnaresi, Gloria Cinquegrani, Raffaella Aldigeri, Francesco Balzamà, Cristina Branchi, Mario Lauria, Riccardo C. Bonadonna, Luca Marchetti, Alessandra Dei Cas

## Abstract

**Background:** Atherosclerosis is increasingly recognized as a chronic immunometabolic disorder involving complex interactions between circulating immune cells, metabolic factors, and the vascular wall. Carotid intima–media thickness (IMT) is widely used as a surrogate marker of subclinical atherosclerosis. Peripheral blood mononuclear cells (PBMCs) provide a systemic readout of immune transcriptional states, but longitudinal evidence linking PBMC transcriptional profiles to long-term vascular remodeling remains limited.

**Methods:** We analyzed the association between PBMC transcriptomic profiles and carotid IMT in the Barilla Offspring Study, a single-center cohort with long-term follow-up. PBMC transcriptomics and carotid IMT were assessed at baseline in 148 participants, and 101 individuals underwent repeat clinical, vascular, and transcriptomic evaluation at follow-up. Three analytical configurations were examined: baseline cross-sectional, follow-up cross-sectional, and a longitudinal model linking baseline transcriptomic profiles to follow-up IMT. Following a comparison of state-of-the-art machine learning regression algorithms and an innovative rank-based method, the most predictive transcriptomic signature from each analytical configuration was used for downstream functional enrichment and network analyses.

**Results:** The rank-based regression method showed the best performance across all analytical configurations. Cross-sectional analyses at both time points consistently revealed enrichment of immune-related pathways, including leukocyte activation, antigen presentation, and receptor-mediated signaling. In contrast, the longitudinal transcriptomic signature was enriched for pathways related to metabolic regulation, redox processes, and cellular structural organization. Despite limited overlap at the single-gene level, functional similarity analysis demonstrated convergence toward shared immunometabolic pathways associated with vascular remodeling.

**Conclusions:** PBMC transcriptional profiles are associated with subclinical vascular remodeling both cross-sectionally and over long-term follow-up, suggesting that systemic immune transcriptional states may contribute to vascular aging.

## Introduction

Cardiovascular disease (CVD) remains the leading cause of global morbidity and mortality, with clinical events largely attributable to the progressive evolution and destabilization of atherosclerotic lesions. Atherosclerosis is currently understood as a chronic immunometabolic disorder of the arterial wall, driven by the complex interactions among genetic susceptibility, environmental exposures, metabolic factors, and immune responses [1].

Although conventional cardiovascular (CV) risk factors explain a substantial proportion of population-level risk, they do not fully account for the molecular and cellular processes that initiate atherogenesis and govern the long-term evolution of vascular lesions during their prolonged subclinical phase [2].

Carotid intima–media thickness (IMT) is widely used as a non-invasive surrogate measure of subclinical atherosclerosis and has been independently and consistently associated with incident CVD [3]. Nonetheless, individuals with comparable traditional risk profiles frequently exhibit markedly heterogeneous longitudinal trajectories of IMT progression, suggesting that unmeasured biological determinants modulate arterial wall remodeling beyond established clinical predictors [4]. Accumulating evidence indicates that circulating immune cells play a central role in atherogenesis [5]. Monocytes and lymphocytes actively participate in endothelial dysfunction, plaque formation, and lesion progression through cytokine production, cell–cell interactions and metabolic reprogramming within the vascular microenvironment [6,7]. In parallel, the framework of immunometabolism has highlighted how systemic metabolic and inflammatory signals shape immune cell function and transcriptional programs [8–10].

Peripheral blood mononuclear cells (PBMCs), which express a substantial proportion of the human transcriptome and are continuously exposed to endogenous and environmental cues [11,12], provide a dynamic readout of systemic immunometabolic processes. Their role as a systemic integrator of metabolic and inflammatory signals makes them an ideal substrate for identifying early signatures of vascular damage.

Cross-sectional studies have linked PBMC gene expression profiles to dietary patterns and to metabolic perturbations in healthy as well as obese and insulin resistant individuals [13–17]. However, available evidence is largely cross-sectional and robust longitudinal data examining whether baseline PBMC transcriptomic signatures anticipate long-term structural vascular changes are lacking. Prospective investigations integrating immune transcriptomics with subclinical vascular phenotyping over extended follow-up periods remain exceedingly scarce.

In a cohort of deeply phenotyped healthy individuals enrolled 18 years ago in the Barilla Offspring Study [18,19], we previously identified, using a rank-based classification approach, a reproducible PBMC gene expression signature associated with insulin resistance, which also discriminated pathological from physiological states in independent cohorts [9].

This cohort is uniquely characterized by comprehensive metabolic and clinical phenotyping at baseline, high-resolution transcriptomic profiling, and the subsequent availability of standardized carotid IMT measurements after nearly two decades of follow-up, thereby enabling a rare longitudinal systems-biology evaluation of immune transcriptional states and vascular remodeling. Taking advantage of this longitudinal design, we investigated the relationship between PBMC transcriptomic signatures and carotid IMT through cross-sectional analyses performed at each time point, together with a prospective analysis evaluating whether baseline transcriptional profiles predicted IMT at the 18-year follow-up. We then assessed predictive performance and explored the associated biological pathways to gain insight into molecular mechanisms potentially involved in long-term vascular remodeling. Because this study spans nearly two decades—a period characterized by substantial advances in biotechnological precision [20,21], temporal changes in experimental technologies were considered. In particular, the transition from microarray-based transcriptomics at baseline to high-depth RNA sequencing at follow-up, together with improvements in carotid ultrasound imaging for IMT assessment [22,23], introduced technological heterogeneity that required methodological adjustment. Because these shifts preclude direct point-by-point comparison of raw signal intensities [24,25], we adopted three complementary analytical strategies: (i) baseline cross-sectional analysis evaluating the association between baseline transcriptomic profiles and baseline IMT, (ii) follow-up cross-sectional analysis evaluating the association between follow-up RNA-seq profiles and the corresponding IMT measurements and (iii) a longitudinal prognostic analysis assessing the capacity of baseline transcriptomic signatures to predict vascular remodeling at 18 years.

## Methods

### Study design and population

This study represents the 18-year follow-up of the Barilla Offspring Study, a single-centre observational cohort established in 2006–2007 (T0) [18,19]. At baseline, 148 healthy adults - recruited among the offspring of participants in the Barilla Factory Study - underwent comprehensive clinical and metabolic characterization, including PBMC transcriptomic profiling by microarray analysis [9].

Of the 148 baseline participants, 101 (68.2%) were re-evaluated at the 18-year follow-up in 2024 (T1). Among the 47 individuals not reassessed, 11 (7.4%) declined participation, one had died, and the remaining were lost to follow-up due to unsuccessful contact attempts or relocation. Inclusion criteria were participation in the baseline study and availability of transcriptomic data. Ongoing pregnancy was among the exclusion criteria. The study was registered at ClinicalTrials.gov (identifier: NCT06876818) and approved by the local Institutional Review Board (Comitato Etico di Area Vasta Emilia Nord; Protocol 45543, 14 November 2023). The study was conducted in accordance with the Declaration of Helsinki, and all participants provided written informed consent prior to study entry.

All clinical, metabolic and vascular assessments performed at baseline were repeated at follow-up. This included age, sex, family history of diabetes and cardiovascular disease (CVD), smoking habits, body mass index (BMI), waist circumference, systolic and diastolic blood pressure, heart rate, and medical history. Venous blood samples were collected after an overnight fast for standard biochemical analyses and peripheral blood mononuclear cell (PBMC) isolation. Oral glucose tolerance testing (OGTT) and carotid ultrasound examination for intima–media thickness (IMT) assessment was performed at both time points.

### Intima-media thickness assessment

In the Barilla Offspring baseline study (T0), carotid intima–media thickness (IMT) was assessed by high-resolution B-mode ultrasound using a linear phased-array multifrequency transducer (7–10 MHz), as previously described [26]. Images were acquired at end diastole using electrocardiographic gating to minimize variability related to carotid pulsatility. IMT measurements were obtained at the distal common carotid artery, carotid bifurcation, and proximal internal carotid artery. Measurements were performed on both near and far walls and analyzed offline by a single experienced operator using a semiautomated edge-detection system (Carotid Analyzer; Medical Imaging Applications LLC, IA, USA).

At follow-up (T1), carotid IMT was assessed using a linear multifrequency transducer (7–10 MHz) mounted on a Logiq E10 ultrasound system. In selected individuals with limited acoustic windows (e.g., short or thick neck), an additional curvilinear transducer (3.5–5 MHz) was used to improve visualization of deeper vascular structures. Examinations were performed with participants in the supine position with the neck extended. Images were obtained from three carotid segments: the distal common carotid artery (last centimeter before bifurcation), the carotid bifurcation, and the proximal internal carotid artery (first centimeter after bifurcation).

IMT was measured on both near and far walls at the distal common carotid artery over a 10 mm segment located at least 5 mm below the bifurcation, and on the far wall at the carotid bifurcation and proximal internal carotid artery. Mean IMT was calculated as the average of all IMT measurements obtained across the examined segments, whereas maximum IMT was defined as the maximum IMT value recorded in the same segments. Measurements were obtained using an automated software system that calculates both parameters.

Due to technological advancements over the 18-year interval - including differences in ultrasound equipment, measurement software, sonographer, and the absence of electrocardiographic gating at follow-up - absolute IMT values obtained at the two time points were not directly compared.

### Measurement of glucose and insulin concentration

Plasma glucose concentrations were measured using an EKF Biosen C-line analyzer (EKF-diagnostic GmbH, Barleben, Germany) based on an enzymatic amperometric method with immobilized glucose oxidase, according to the manufacturer’s instructions [27].

Plasma insulin concentrations were determined using a commercially available human insulin ELISA kit (10-1113-01, Mercodia, Uppsala, Sweden), according to the manufacturer’s instructions. Absorbance was measured at 450 nm using a Multiskan™ FC microplate reader (Thermo Fisher Scientific, Waltham, MA, USA).

### Gene expression profiling of PBMCs

#### PBMCs isolation

PBMCs were isolated by density-gradient centrifugation using Lympholyte-H Cell Separation Medium (CL5020-R, CEDERLANE, Burlington, ON, Canada). Cells were washed twice with Hanks’ Balanced Salt Solution (HBSS; L0611, Dutscher, Bernolsheim, France) and incubated with ammonium chloride potassium (ACK) lysis buffer for 10 minutes on ice to remove residual red blood cells, followed by an additional wash in HBSS. After cell counting, 5 × 10[PBMCs were lysed in RLT buffer and stored at −80 °C until RNA extraction.

#### RNA extraction

Total RNA was extracted using the RNeasy Mini Kit (74106, Qiagen, Hilden, Germany) according to the manufacturer’s protocol. Cells were lysed in 350 µL of RLT buffer supplemented with β-mercaptoethanol and homogenized using a QIAshredder spin column (79656, Qiagen). Genomic DNA contamination was removed by RNase-free DNase treatment (79254, Qiagen). RNA concentration and purity were assessed using a NanoDrop One spectrophotometer (Thermo Fisher Scientific, Waltham, MA, USA), and RNA integrity was evaluated with a 4200 TapeStation system (Agilent Technologies, Santa Clara, CA, USA). Only samples with an RNA integrity number (RIN) > 8 were included in downstream sequencing analyses.

#### RNA sequencing and transcriptomic analysis

RNA sequencing was carried out by Eurofins Genomics (Ebersberg, Germany). Total RNA was subjected to ribosomal RNA and globin mRNA depletion, followed by fragmentation and the preparation of strand-specific cDNA libraries using random priming. Libraries were generated through adapter ligation and PCR amplification and subsequently sequenced on an Illumina NovaSeq platform (2 × 150 bp paired-end reads), with a guaranteed sequencing depth of 30 million read pairs per sample (±3%). Raw sequencing data were delivered as FASTQ files for downstream bioinformatic analyses.

#### Bioinformatic and transcriptomic analysis

Transcriptomic data from the baseline cohort were retrieved from the Gene Expression Omnibus [28] (GEO) under accession number GSE87005. Microarray data (two-channel from the Agilent platform) were then processed using the R Bioconductor package limma [29]. Background correction was performed using the normexp method, followed by within-array and between-array normalization using the loess and a quantile method, respectively. To ensure gene-level consistency, probes targeting the same gene were collapsed to a single value using their mean expression. Finally, probe identifiers were mapped to official gene symbols via the gconvert function [30].

For RNA-seq data, raw paired-end FASTQ files were aligned to the GRCh38 human reference genome using STAR [31] (v2.7.11b). Alignment was conducted using the option “two-pass mode Basic” to optimize splice junction discovery, guided by the GENCODE v48 primary assembly annotation. Gene level quantification was performed directly during alignment via the GeneCounts parameter. Raw counts were normalized to Transcripts per Million [32] (TPM) to account for gene length and library size. The resulting values were log2-transformed to stabilize variance. Ensembl IDs were mapped to gene symbols using GENCODE v48 metadata whenever possible. In instances where multiple IDs mapped to a single symbol, the mean expression value was utilized.

To minimize the impact of non-biological variance and potential confounding factors, we performed covariate correction on both the transcriptomic data and the clinical target variables [24,33]. Age and sex were identified as primary covariates for adjustment. To do so, we fitted an Ordinary Least Squares regression model [34] for each gene and IMT and performed downstream analysis on the residualized matrices.

To model the relationship between transcriptomic profiles and mean IMT we defined three distinct analytical configurations: (i) Baseline: baseline transcriptome paired with baseline IMT, (ii) Follow-Up: Follow-Up transcriptome paired with follow-up IMT and (iii) Prognostic: baseline transcriptome paired with follow-up IMT. The baseline model included all subjects with available baseline transcriptomic and IMT data (n = 148), whereas the follow-up and prognostic models were restricted to participants with measurements available at both time points (n = 101).

We compared three regression methods: LASSO [35], Random Forest Regression [36], and a modified rank-based classification algorithm, initially proposed in [37,38], adapted for regression tasks. This method, extended with a global optimizer to learn the transcriptomic signatures providing the best regression performance, has already been successfully applied in its classification version to the baseline cohort dataset [9] and demonstrated its efficacy during the SBV IMPROVER Diagnostic Signature Challenge [39] and in several other studies [40–45]. The method summarizes each sample’s characteristics through a rank-based gene signature and performs an all-to-all signature comparison by means of a distance metric based on weighted enrichment score (ES) [46]. These comparisons are compiled in a distance matrix, which is then used to predict a subject’s mean IMT as the weighted average of the neighbours. A transcriptomic signature is finally returned for each analytical configuration, collecting all the genes that appear in at least one subject-specific signature. Model performance computed by the three considered regression algorithms was evaluated using a 10-fold cross-validation scheme, and the method yielding the highest Ratio of Performance to Deviation (RPD) was selected for feature extraction. The signatures identified in the three analytical configurations (*i.e.,* Baseline, Follow-Up, and Prognostic) by the top-performing method were used downstream for functional analysis.

Functional enrichment of the prioritized gene signatures was conducted using the STRING database [47]. The analysis initially incorporated a primary shell of 10 functional partners, expanded to 20 if the Protein-Protein Interaction (PPI) enrichment p-value exceeded 0.05, to capture broader pathway associations and ensure statistical robustness. We evaluated the convergence of the three analytical configurations by identifying shared genes and calculating functional semantic similarity. Semantic similarity was assessed within the Gene Ontology (GO) Biological Process domain [48] using the GOATOOLS library [49] and human-specific annotations. We calculated the similarity between GO terms using Resnik’s Information Content (IC) [50] method, which identifies the IC of the Most Informative Common Ancestor (MICA). Term-level scores were aggregated into a gene-set-level similarity score using the Best-Match Average (BMA) algorithm, which provides a symmetric average of the maximum similarities between sets.

Finally, we constructed a functional interaction network (STRING [47], confidence score >0.4). Within this network, “hub genes” were identified based on degree centrality (degree >= 3), which indicates the total number of edges in the network involving them. These high-connectivity nodes were prioritized as the central biological drivers of the shared transcriptomic response.

### Statistics

For clinical data continuous variables are presented as means and standard deviation or median and interquartile range (IQR) whereas categorical data are presented as numbers (percentages).

Normality of continuous variables was verified with one-way Kolmogorov–Smirnov test and non-normal variables were log-transformed before analysis with parametric tests. Comparison between cohorts was performed by means of paired t- test or Wilcoxon, as appropriate, including only participants with measurements available at both time points (n = 101) or χ2 test for categorical variables. Baseline descriptive statistics, however, include all participants with available data at baseline (n = 148). All tests were two-sided with p<0.05 considered statistically significant. Statistical analyses were performed using SPSS version 30.0 (IBM Statistics).

## Results

### Clinical characteristics of study population

Baseline and 18-year follow-up characteristics of the participants are summarized in Table 1. The cohort in follow-up (N=101) was not different in the key characteristics from the one that dropped out (N=47), confirming that the follow-up cohort remained broadly representative of the original population. (data not shown). Over time, the cohort showed the expected age-related shift toward a less favourable cardiometabolic profile, characterized by increases in adiposity measures, blood pressure, fasting glucose, triglycerides, and selected liver enzymes.

**Table 1.** Demographic and laboratory characteristics of study cohort at two time points.

|  | **Baseline cohort T0**<br>**N=148** | **Follow-up cohort T1**<br>**N=101** | **p-value** |
| --- | --- | --- | --- |
| **Age(yrs)** | 37.0±8.0 | 54.1±7.3 | <0.001 |
| **Male sex** | 82(55) | 53(51) | 1.00 |
| **Height(cm)** | 172±9 | 170±10 | 1.00 |
| **Weight(kg)** | 71.4±13.4 | 73.77±14.85 | <0.001 |
| **BMI (kg/m<sup>2</sup>)** | 23.9(21.8-26.1) | 25.3(23.0-27.8) | <0.001 |
| **Waist circumference(cm)** | 87.2±9.0 | 90.1±12.8 | <0.001 |
| **Systolic pressure(mmHg)** | 115.6±14.5 | 124.7±16.0 | <0.001 |
| **Diastolic pressure(mmHg)** | 74.7±11.8 | 82.4±10.9 | <0.001 |
| **Cardiac frequency(bpm)** | na | 70±11 |  |
| **Current smoking** | 37(25) | 20(19.8) | <0.001 |
| **Pack_yr** | 3.2±7.0 | 13.2±13.8 | 0.014 |
| **White Blood Cell count**<br>**(x10<sup>9</sup>/L)** | 5.88±1.32 | 5.91±1.54 | 0.81 |
| **Neutrophil count (x10<sup>9</sup>/L)** | 3.30(2.70-3.90) | 3.26(2.64-3.92) | 0.95 |
| **Lymphocyte count(x10<sup>9</sup>/L)** | 1.80(1.60-2.12) | 1.75(1.49-2.14) | 0.26 |
| **Monocyte count(x10<sup>9</sup>/L)** | 0.40(0.30-0.50) | 0.44(0.33-0.53) | <0.001 |
| **Monocyte-to-Lymphocyte**<br>**Ratio** | 0.21(0.17-0.26) | 0.25(0.19-0.29) | <0.001 |
| **Neutrophil-to-Lymphocyte**<br>**Ratio** | 1.75(1.41-2.20) | 1.77(1.49-2.26) | 0.81 |
| **Platelet-to-Lymphocyte Ratio** | 136.5(111,7-166.29) | 140.04±45.49 | 0.12 |
| **Haemoglobin(g/dL)** | 14.27±1.63 | 14.29±1.26 | 0.87 |
| **Platelet count(x10<sup>9</sup>/L)** | 255.7±55.7 | 242.2±52.4 | <0.001 |
| **HbA<sub>1c</sub>(mmol/mol)** | na | 34.0(32.0-37.0) |  |
| **FPG (mg/dL)** | 86.5±6.6 | 95.9±10.0 | <0.001 |
| **Total cholesterol(mg/dL)** | 204.58±33.13 | 191.25±36.74 | 0.001 |
| **HDL cholesterol(mg/dL)** | 59.80±14.97 | 62.24±14.53 | 0.02 |
| **LDL cholesterol(mg/dL)** | 129.73±30.62 | 110.87±31.92 | <0.001 |
| **Triglycerides(mg/dL)** | 59.0(47.0-92.0) | 79.0(64.0-98.0) | <0.001 |
| **GGT(U/L)** | 16.0(13.0-23.0) | 21.0(15.0-31.0) | <0.001 |
| **AST(U/L)** | 22.0(19.0-26.0) | 23.0(20.0-28.0) | 0.003 |
| **ALT(U/L)** | 20.0(16.0-27.0) | 22.0(16.0-28.0) | 0.37 |
| **Creatinine(mg/dL)** | 0.92±0.16 | 0.83±0.18 | <0.001 |
| **eGFR(ml/min/1.73m<sup>2</sup>)** | 95.7(85.3-107.2) | 94.0(84.0-102.0) | <0.001 |
| **IMT mean (mm)** | 0.679(0.640-0.725) | 0.678(0.620-0.740) | 0.06 |
| **IMT max (mm)** | 0.830(0.750-0.895) | 1.060(0.940-1.160) | <0.001 |
| **Carotid plaques** | 4(2.7%) | 33(32.7%) | <0.001 |

Data are presented as mean ± standard deviation for normally distributed variables and median (25°-75°) for non-normally distributed variables. Categorical variables are expressed as frequencies (%). P-values from paired t-test or Wilcoxon, as appropriate. BMI: body mass index; FPG: fasting plasma glucose; GGT: gamma-glutamyl transferase; AST: aspartate aminotransferase; ALT: Alanine Aminotransferase; eGFR: estimated Glomerular Filtration Rate; IMT: intima media thickness.

In contrast, total and LDL cholesterol levels were significantly lower at follow-up, whereas HDL cholesterol was modestly higher. At follow-up, 29 participants (28.7%) were receiving lipid-lowering therapy (Table 2), a factor that should be considered when interpreting lipid profile changes. Overall white blood cell counts remained unchanged; however, the monocyte-to-lymphocyte ratio increased significantly, indicating a relative redistribution within circulating leukocyte subsets despite stable total leukocyte counts.

**Table 2.**
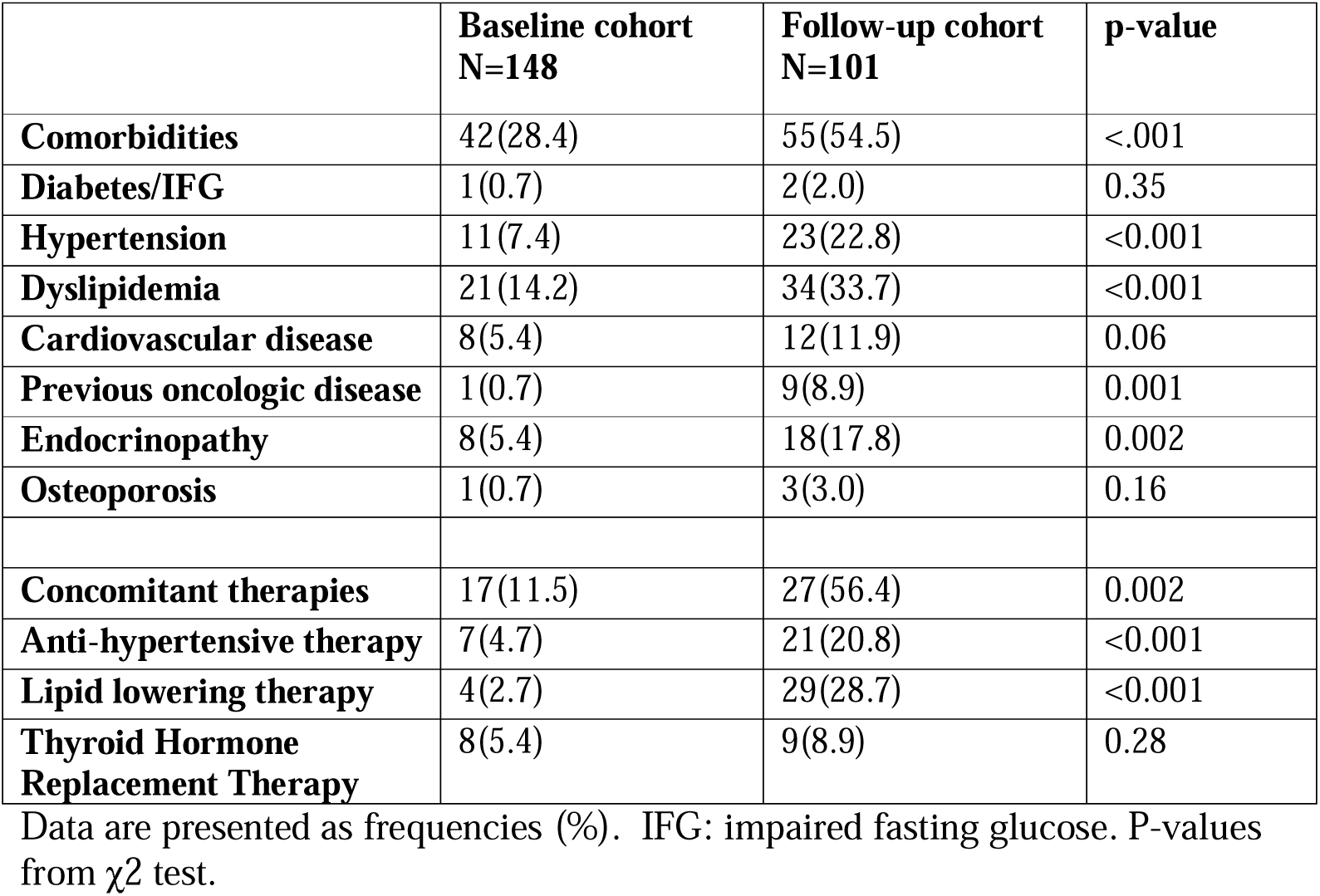
Concomitant diseases and therapies in the follow-up cohort.

Of note, mean carotid IMT values were similar at baseline and follow-up (Table 1), whereas maximum IMT values and carotid plaque prevalence increased significantly, consistent with progression of structural atherosclerotic burden over the 18-year period. However, direct longitudinal comparison of absolute IMT measurements should be interpreted with caution because of differences in ultrasound methodology between the two examinations. Given that mean IMT provides a more reproducible estimate of diffuse arterial wall thickening and is less influenced by focal plaque formation than maximum IMT, mean IMT was used as the primary vascular phenotype in subsequent transcriptomic analyses, in line with previous epidemiological studies of carotid IMT [51].

### Transcriptomics

Transcriptomic profiles from the baseline and follow-up cohorts were normalized (Supplementary Figure S1, S2, S3, S4 and S5) and corrected for age and sex following the procedure outlined in Methods, subsection “Bioinformatic and transcriptomic analysis”. Principal component analysis confirmed that the variance attributable to age and sex, which was minimal at baseline, was successfully mitigated across all three data configurations (Supplementary Figure S6 and S7). The residualized matrices after covariate correction served as the standardized input for all downstream biomarker discovery.

We evaluated three machine learning algorithms: LASSO, Random Forest Regression, and an innovative Rank-Based Regression Method, across the three analytical configurations (Baseline, Follow-up, and Prognostic). As illustrated in Figure 1, the rank-based approach consistently outperformed its counterparts. Specifically, the rank-based algorithm achieved a consistently higher Ratio of Performance to Deviation (RPD) across all analytical configurations: (i) Baseline: RPD 1.38 (vs. 1.07 and 0.99), (ii), Follow-up: RPD 1.26 (vs. 0.97 and 0.97) and (iii) Prognostic: RPD 1.38 (vs. 0.98 and 0.96). Consequently, the rank-based algorithm was selected as the primary method for biomarker extraction.

**Figure 1.**
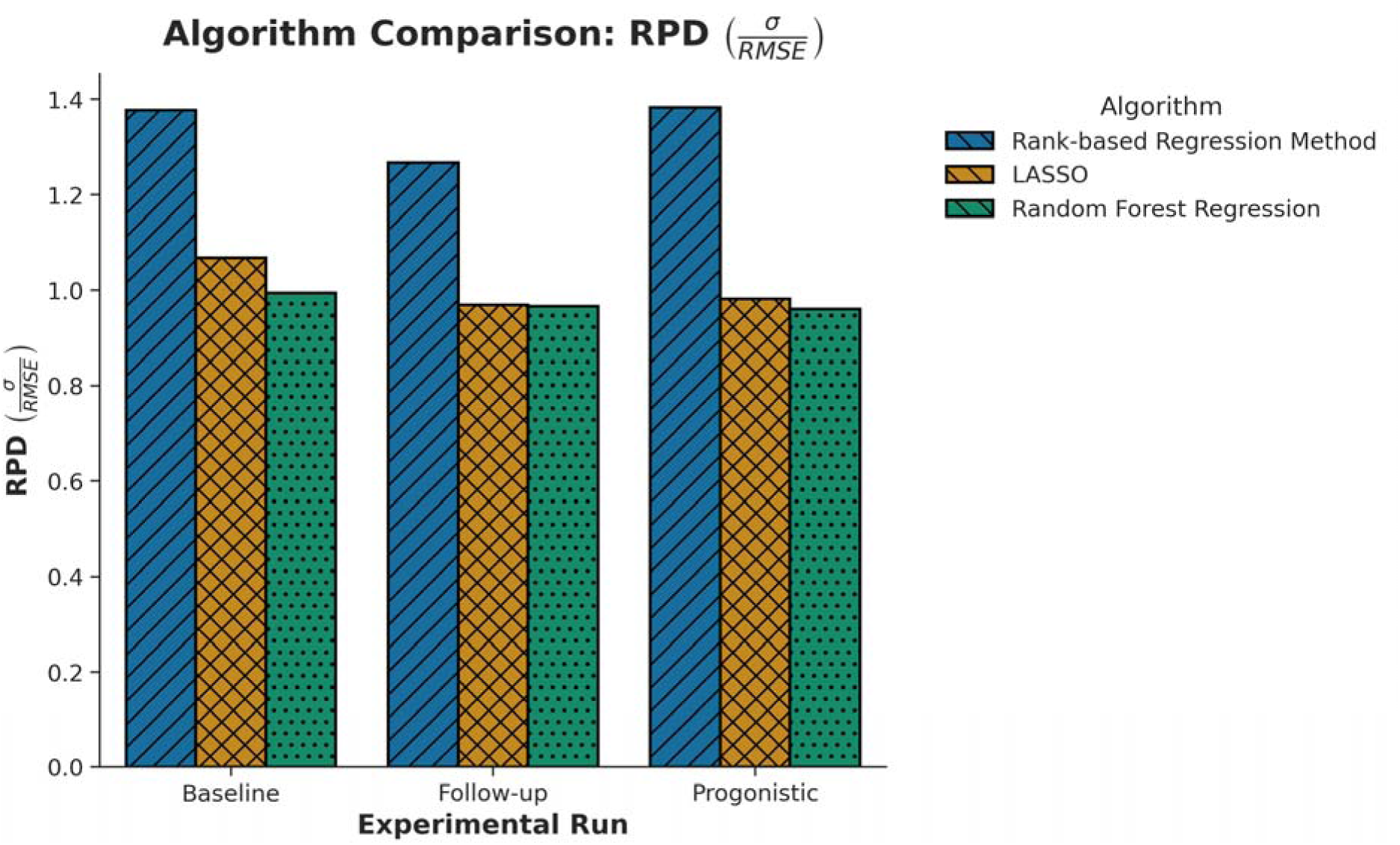
Predictive performance across experimental configurations. The bar plot illustrates the Ratio of Performace to Deviation (RPD) for the three evaluated machine learning models: the Rank-based method (blue), LASSO (orange), and Random Forest Regression (green). The rank-based algorithm consistently demonstrated better predictive power across the Baseline (RPD = 1.38), Follow-up (RPD = 1.26), and Prognostic (RPD = 1.38) configurations, justifying its selection for downstream biomarker extraction.

The biomarker discovery pipeline identified distinct gene sets for each study configuration, available in Supplementary Data (Supplementary File S1): a cross-sectional baseline signature of 114 genes (permutation test p-value: 0.035), a cross-sectional follow-up signature of 187 genes (permutation test p-value: 0.0414), and a longitudinal prognostic signature of 69 genes, which demonstrated the ability to predict 18-year mean IMT progression using only the baseline (2006) transcriptomic profile (permutation p-value: 0.0094). Despite the large temporal and technological gap between the baseline and follow-up, the two transcriptomic signatures shared the WNK4 gene, a known regulator of blood pressure and electrolyte balance.

The biological relevance of these signatures was assessed by a functional enrichment analysis via the STRING database. Baseline (Figure 2) and follow-up (Figure 3) enrichments revealed a robust signal related to immune activation and receptor signaling. Specifically, the baseline signature was significantly enriched for the regulation of leukocyte proliferation, while the follow-up was dominated by T-cell receptor signaling and antigen receptor-mediated pathways.

**Figure 2.**
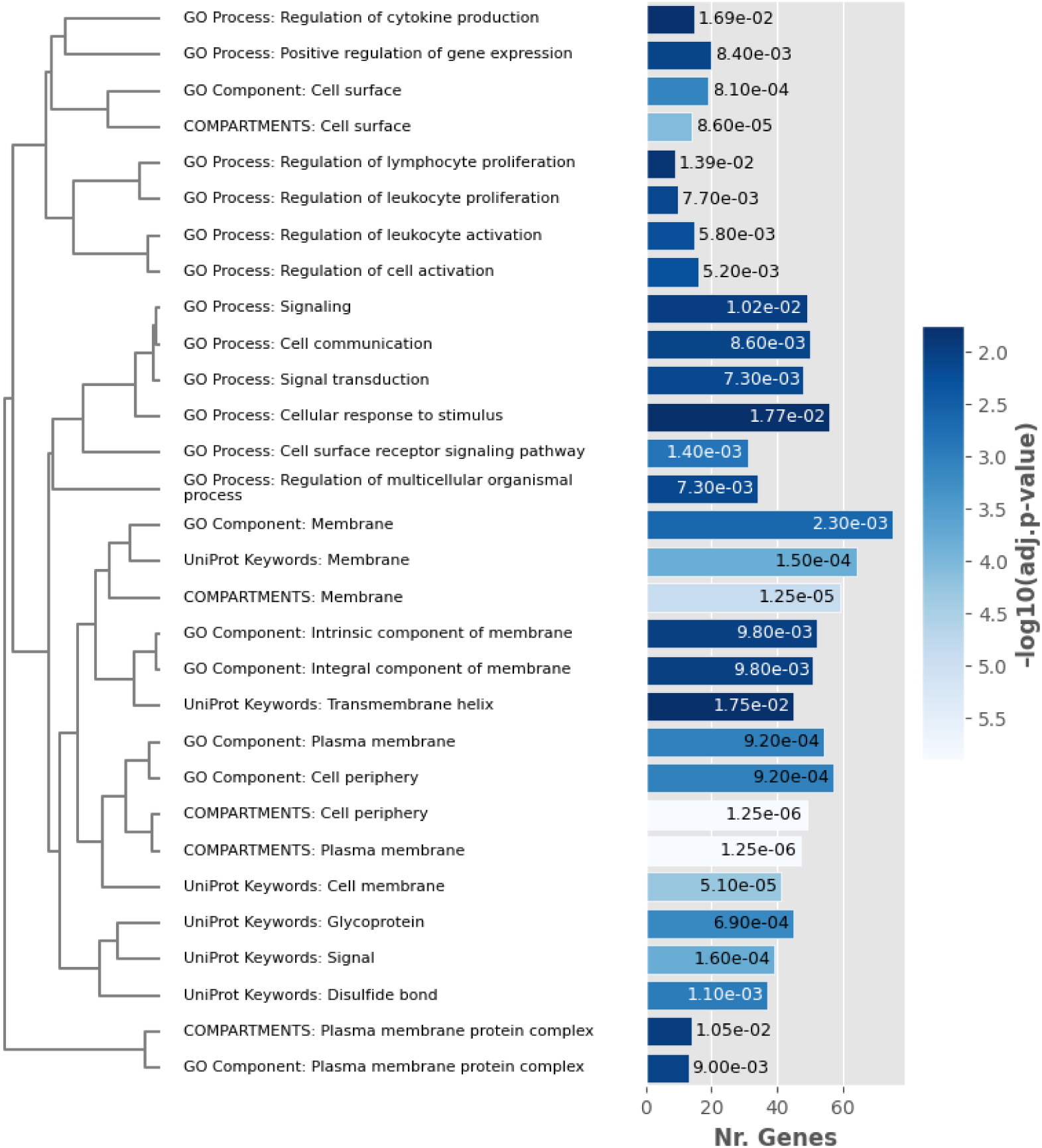
Functional enrichment analysis of the Baseline (2006) transcriptomic signature. The barplot illustrates the top 30 biological processes and pathways associated with contemporary mean IMT at baseline (10 interactors). Enrichment is predominantly characterized by immune-mediated signaling and leukocyte regulation, reflecting the active inflammatory state of the vasculature at the time of sampling.

**Figure 3.**
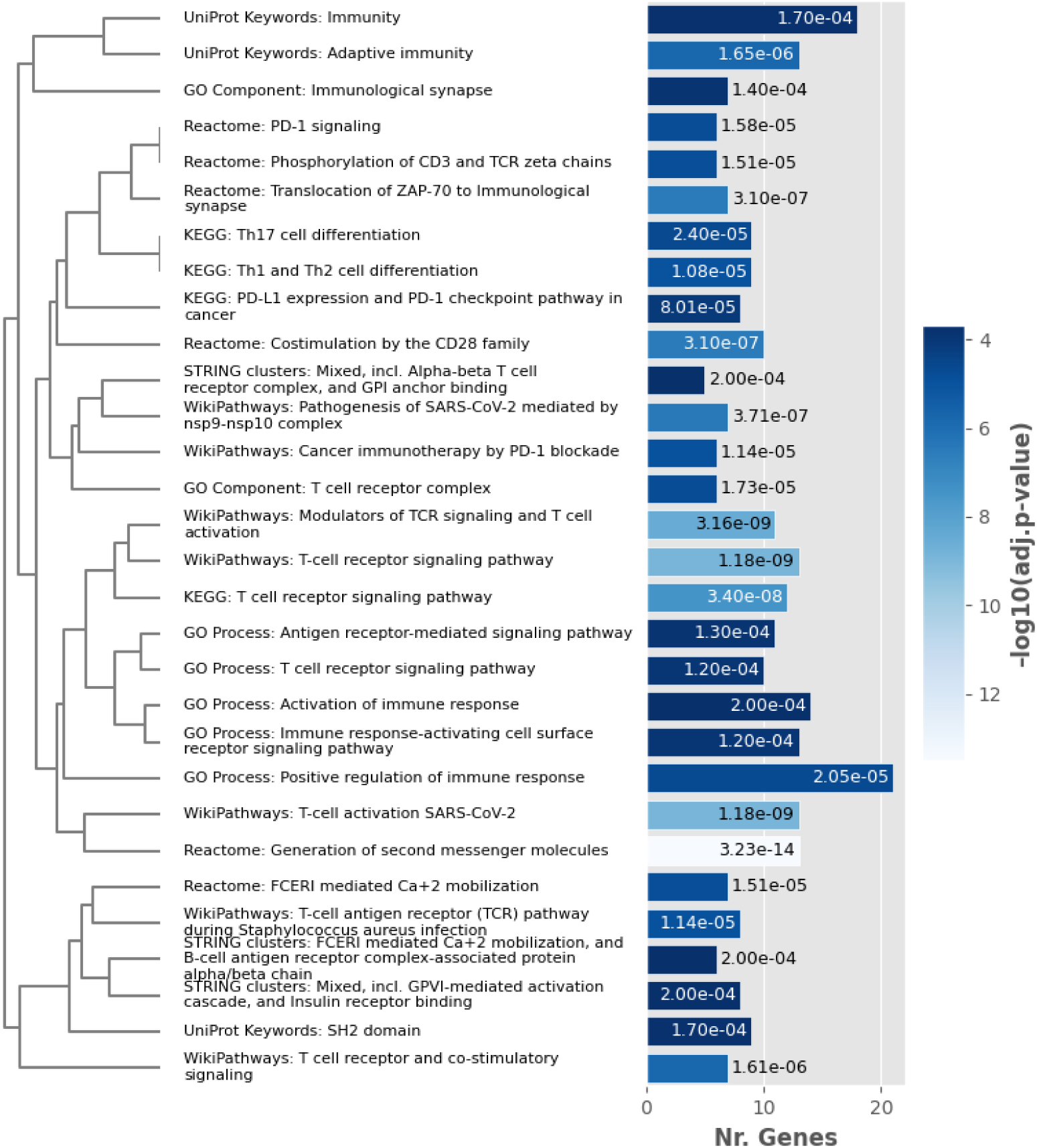
Functional enrichment analysis of the follow-up transcriptomic signature. The barplot illustrates the top 30 biological processes and pathways associated with contemporary IMT at follow-up (10 interactors). Like the baseline results, the signature is heavily weighted toward adaptive immune responses and receptor-mediated signaling.

In contrast, the prognostic signature (Figure 4) was instead characterized by metabolic regulation and structural integrity. Key enriched terms included folate biosynthesis, D-xylose metabolism, and pathways associated with diabetic cataract, reflecting systemic chronic oxidative stress.

**Figure 4.**
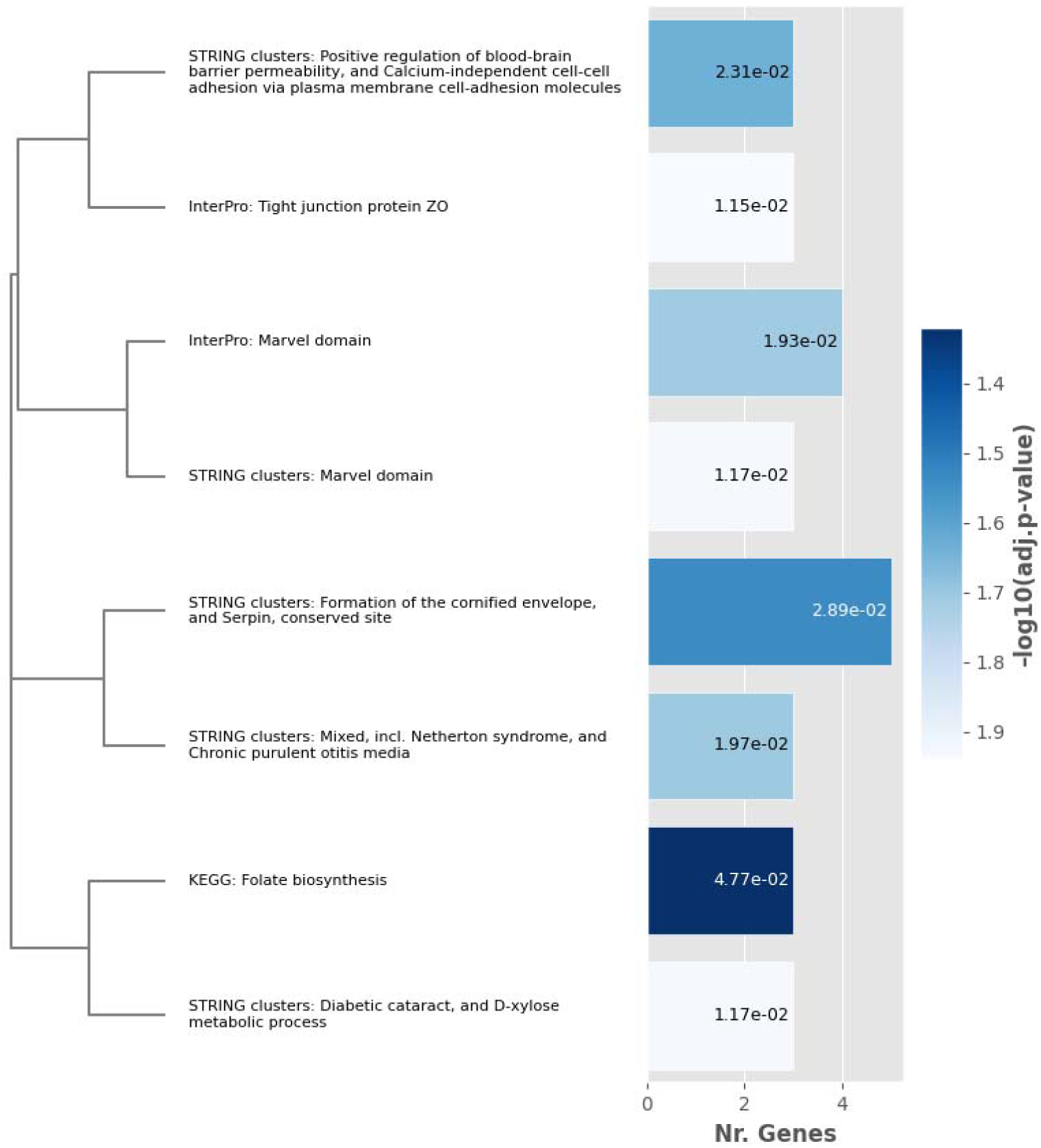
Functional enrichment analysis of the prognostic transcriptomic signature. The barplot illustrates the top 30 biological processes and pathways associated with long-term IMT progression (20 interactors). This signature is characterized by metabolic pathways and structural integrity markers, suggesting that early-stage metabolic stress and cellular apposition are primary drivers of future vascular remodeling.

Furthermore, the presence of MARVEL domain proteins suggests long-term remodeling of membrane apposition and tight junction regulation, potentially impacting vascular endothelial stability.

Despite the minimal direct gene overlap between the transcriptomic signatures, semantic similarity analysis revealed substantial functional convergence between the baseline and follow-up signatures (Resnik-BMA: 4.26). These findings indicate that the transition from microarray to RNA-seq technologies and the long temporal gap did not substantially affect the identification of shared immunometabolic pathways.

Interestingly, the prognostic transcriptomic signature exhibited a more divergent functional profile relative to the baseline signature (Resnik-BMA: 3.76). This lower similarity may reflect distinct biological processes associated with current vascular state and long-term remodeling propensity: while current vascular state is primarily defined by active inflammatory signaling, the underlying propensity for future remodeling is rooted in distinct metabolic and structural foundations.

The biological validity of these associations is further supported by the joint network analysis (Figures 5 and Figure 6). These networks exhibit a “well-mixed” topology, where genes from different discovery cohorts integrate into interconnected functional modules rather than forming isolated, platform-specific clusters. This finding strongly supports the robustness of our analysis against potential technical bias. By assessing the topological centrality within these integrated networks, we identified several high-degree hub genes (degree >= 3). These hubs act as the primary biological mediators, anchoring the shared molecular response across all analytical configurations.

**Figure 5.**
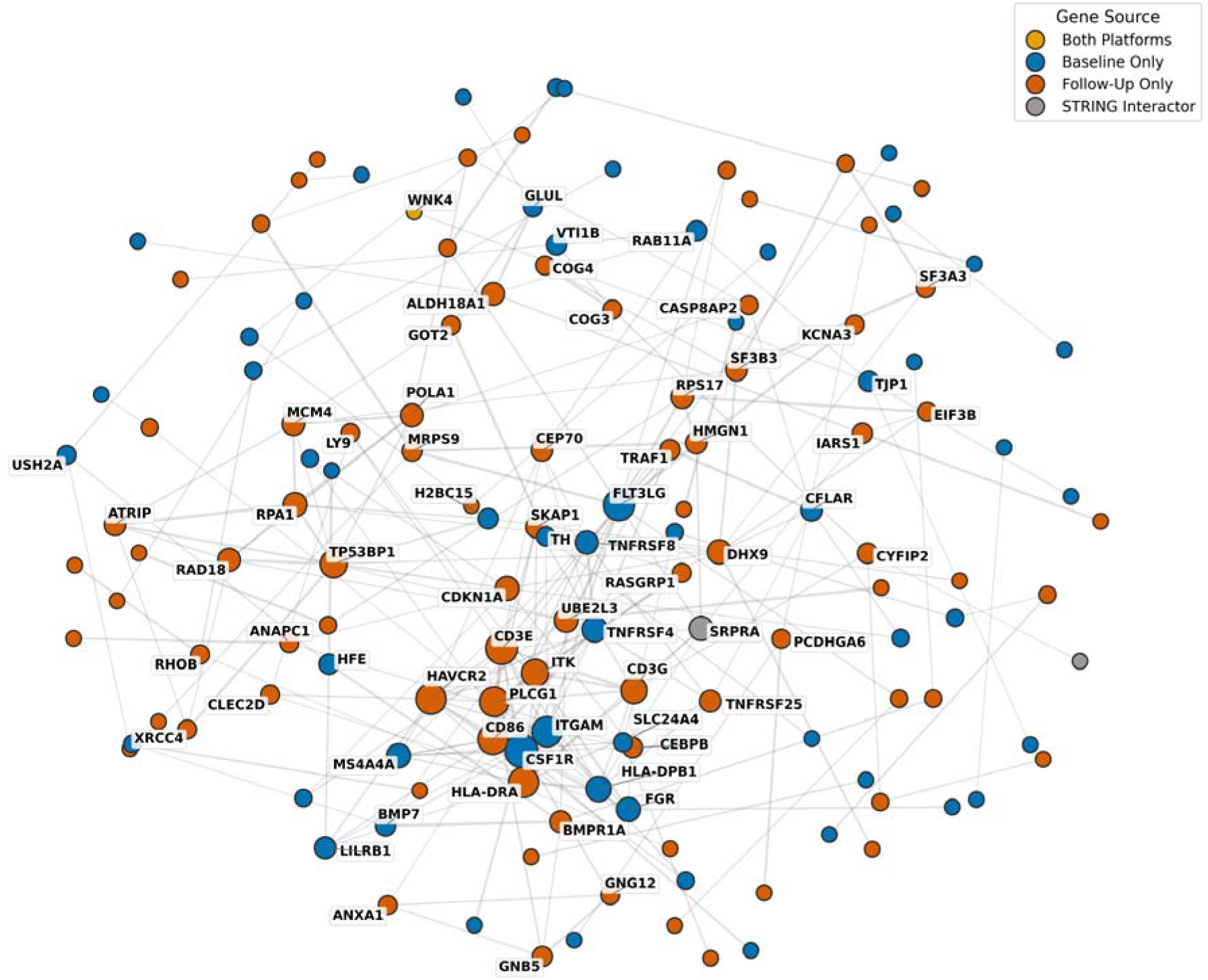
Joint Network Topology and Functional Integration (Baseline vs. Follow-up). The interaction network demonstrates the topological integration of biomarkers identified from the baseline and 18-year follow-up cohorts. Labels identify hub genes (degree >= 3).

**Figure 6.**
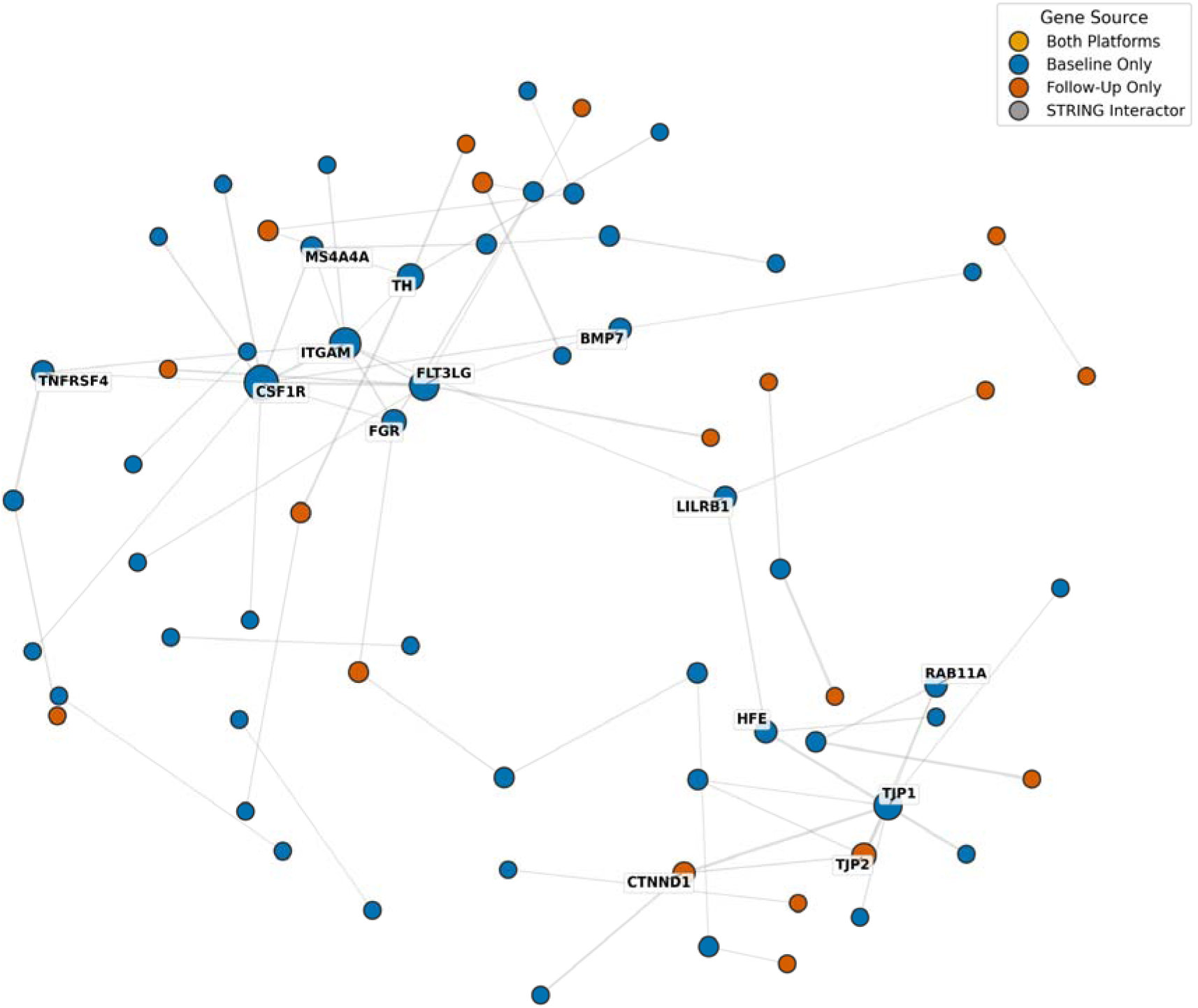
Joint Network Topology and Functional Integration (Baseline vs. Prognostic). The interaction network demonstrates the topological integration of biomarkers identified from the cross-sectional baseline discovery and the longitudinal prognostic model. Labels identify hub genes (degree >= 3).

The identification of these hubs, summarized in Tables 3 and 4, demonstrates that, although specific gene candidates may vary between analytical configurations, the core biological drivers of vascular remodeling remain highly consistent across the baseline and the follow-up analysis. This is particularly evident in the integrated networks where shared immunometabolic signals persist across the 18-year interval. Furthermore, we identified a smaller but relevant overlap between the baseline and the prognostic signatures, suggesting the presence of key biological actors which are not only associated with current mean IMT but also actively participate in the molecular programming of its progression over 18 years.

**Table 3.** Top High-Degree Hub Genes in the Baseline vs. Follow-up Integrated Network. This table lists the most highly connected nodes (degree >= 3) within the merged baseline and follow-up transcriptomic signatures.

| Baseline vs Follow-Up<br>Hub genes | Degree | Baseline vs Follow-Up<br>Hub genes | Degree |
| --- | --- | --- | --- |
| CSF1R | 16 | LILRB1 | 5 |
| CD3E | 15 | GNB5 | 4 |
| FLT3LG | 14 | TJP1 | 4 |
| HLA-DRA | 13 | H2BC15 | 4 |
| HAVCR2 | 13 | MRPS9 | 4 |
| CD86 | 13 | VTI1B | 4 |
| ITGAM | 13 | CYFIP2 | 4 |
| PLCG1 | 12 | RAB11A | 4 |
| ITK | 10 | TRAF1 | 4 |
| TP53BP1 | 10 | CEBPB | 4 |
| CD3G | 9 | BMP7 | 4 |
| HLA-DPB1 | 8 | HFE | 4 |
| TNFRSF4 | 8 | IARS1 | 4 |
| CDKN1A | 7 | GLUL | 3 |
| RPA1 | 7 | USH2A | 3 |
| DHX9 | 7 | GNG12 | 3 |
| UBE2L3 | 7 | RASGRP1 | 3 |
| FGR | 7 | GOT2 | 3 |
| MS4A4A | 7 | TH | 3 |
| SRPRA | 7 | COG3 | 3 |
| ALDH18A1 | 6 | COG4 | 3 |
| RAD18 | 6 | CLEC2D | 3 |
| MCM4 | 6 | LY9 | 3 |
| POLA1 | 6 | ANXA1 | 3 |
| TNFRSF8 | 6 | RHOB | 3 |
| RPS17 | 6 | XRCC4 | 3 |
| SKAP1 | 5 | PCDHGA6 | 3 |
| CFLAR | 5 | SLC24A4 | 3 |
| ATRIP | 5 | CASP8AP2 | 3 |
| HMGN1 | 5 | ANAPC1 | 3 |
| BMPR1A | 5 | SF3A3 | 3 |
| CEP70 | 5 | EIF3B | 3 |
| SF3B3 | 5 | KCNA3 | 3 |
| TNFRSF25 | 5 |  |  |

**Table 4.** Top High-Degree Hub Genes in the Baseline vs. Prognostic Integrated Network. This table lists the most highly connected nodes (degree >= 3) within the merged baseline and prognostic transcriptomic signatures.

| Baseline vs Prognostic Hub genes | Degree |
| --- | --- |
| CSF1R | 10 |
| ITGAM | 8 |
| FLT3LG | 7 |
| TJP1 | 6 |
| TH | 5 |
| TJP2 | 4 |
| FGR | 4 |
| RAB11A | 3 |
| TNFRSF4 | 3 |
| BMP7 | 3 |
| MS4A4A | 3 |
| CTNND1 | 3 |
| LILRB1 | 3 |
| HFE | 3 |

## Discussion

In this longitudinal study integrating immune-cell transcriptomics with comprehensive vascular phenotyping, peripheral blood transcriptional signatures were associated with mean carotid IMT both cross-sectionally and prospectively. The identification of robust transcriptomic signatures requires an analytical framework capable of separating subtle biological signals from high-dimensional noise. Our comparative evaluation identified the rank-based regression method as the best choice for biomarker discovery, consistently yielding higher Ratio of Performance to deviation (RPD) values (1.26-1.38) than LASSO or Random Forest Regression across all analytical configurations. While traditional chemometric standards classify models with an RPD < 1.4 as non-reliable for absolute quantification and require an RPD > 2.0 for individual patient-level diagnostics [52], these strict thresholds assume the high signal-to-noise ratios of physical spectroscopy. In the context of high-dimensional-omics data predicting a multifactorial trait, our algorithm effectively extracted and compiled population-level information into a significant and informative transcriptomic signature, although the achieved RPD values suggest that further methodological refinement and additional data may improve predictive performance. By avoiding the extreme sparsity often seen in penalized methods like LASSO, the rank-based approach retained a sufficiently comprehensive gene set to enable meaningful downstream functional enrichment.

Crucially, as these algorithms were trained on residualized matrices, the extracted genes represent a transcriptomic signal independent of established clinical confounders like age and sex. This methodological rigor ensures that the subsequent biological pathways reflect molecular processes directly involved in vascular remodeling rather than mirroring the existing cardiometabolic risk burden.

Our results provide compelling long-term evidence linking systemic immune regulation to the progression of subclinical vascular remodelling. Across all analytical frameworks, transcriptomic signals consistently converged on pathways related to myeloid activation, antigen presentation, and adaptive immune signalling.

These findings indicate that circulating immune cells may capture molecular processes directly involved in vascular remodeling rather than merely reflecting the established cardiometabolic risk burden.

Atherosclerosis is widely regarded as a chronic immunometabolic disorder arising from continuous interactions among circulating immune cells, metabolic exposures, and the vascular microenvironment. In this context, PBMC transcriptional profiles can be viewed as an integrated systemic readout of immune activation states shaped by long-term environmental and metabolic influences.

Several hub genes emerging from the network analysis correspond to biological mechanisms implicated in the initiation and progression of atherogenesis. In the cross-sectional baseline and follow-up models, protein–protein interaction analysis identified highly connected hub genes predominantly involved in immune-related processes, including CSF1R, CD3E, CD86, and HLA-DRA. These genes highlight coordinated remodeling of innate and adaptive immune pathways associated with IMT when transcriptomic and clinical measurements are obtained at the same time point.

Notably, CSF1R signaling - an essential regulator of monocyte survival and differentiation into tissue macrophages - has been mechanistically linked to lesion development in experimental models. Disruption of the CSF1–CSF1R axis markedly reduces macrophage expansion and atherosclerotic lesion development in murine models [53–55] while increased CSF1R expression characterizes macrophage-rich and advanced human plaques [56]. Complementary to myeloid activation, the involvement of antigen-presenting and T-cell receptor components emerging from enrichment analysis supports a model in which antigen-driven immune responses and myeloid activation operate in concert to sustain arterial wall remodeling. This is consistent with recent *in vivo* studies which, through transcriptomic analysis and Mendelian randomization (MR), have demonstrated a causal association between HLA-DR expression on dendritic cells and CD8 expression on CD8[T cells with atherosclerosis risk [57]. The identification of CD86 and CD3E as hub genes further corroborates the coordinated activation of antigen presentation and adaptive immune responses, supporting the concept that innate and T-cell–mediated immunity synergistically contributes to lesion progression [58,59]. In parallel, ITGAM (CD11b), a key mediator of leukocyte adhesion and transendothelial migration, promotes the early recruitment of inflammatory cells into the arterial wall [60,61]. Notably, these immune signals were already detectable in circulating cells before clinical manifestations of vascular disease became evident, suggesting that systemic immune regulation may precede measurable structural arterial alterations. In addition to these immune signaling networks, the comparison of transcriptomic signatures also revealed individual genes exhibiting notable temporal persistence. The comparison of signature gene lists revealed WNK4 (With-No-Lysine Kinase 4) as one of the few genes shared between cross-sectional baseline and follow-up models. Although WNK4 did not emerge as a network hub, its persistence across nearly two decades suggests a stable association with IMT, independent of the dynamic reorganization of immune-related networks. WNK4 is a serine/threonine kinase traditionally known for regulating renal electrolyte homeostasis and blood pressure via the modulation of sodium-chloride reabsorption [62,63].

Beyond its classical renal functions, WNK4 has recently been implicated in immune responses, including lipopolysaccharide-induced macrophage activation, suggesting a broader role in systemic inflammatory processes [64]. In our study, the identification of WNK4 as a persistent marker suggests that this kinase may represent a molecular link between long-term electrolyte balance, blood pressure regulation, and immune-mediated vascular remodeling. These findings indicate that while most immune pathways undergo longitudinal modulation, certain signals such as WNK4 may reflect relatively stable systemic regulatory mechanisms contributing to vascular aging.

The functional consistency observed over nearly two decades is further supported by the semantic similarity analysis. By focusing on the functional relationships among genes rather than on individual gene identities, we show that the biological processes associated with vascular remodeling remain largely preserved. Thus, although specific transcriptomic signatures may vary with aging or technological differences between platforms, the core pathway architecture underlying vascular remodeling appears to remain conserved.

A central insight from this study emerges from the comparison between cross-sectional and prospective analytical models. In the cross-sectional configuration, pathway enrichment analysis revealed predominant activation of immune-related processes, including regulation of leukocyte proliferation, receptor-mediated signaling, MHC class Ib protein complex binding, GPI-anchor binding, and type II hypersensitivity responses. These findings are consistent with the hub gene profile, dominated by immune and myeloid regulators such as CSF1R, ITGAM, LILRB1, and TNFRSF4, collectively indicating enhanced leukocyte activation, antigen presentation, and inflammatory signaling [65]. Specifically, LILRB1 and the TNFRSF4 (OX40) co-stimulatory pathway have been implicated in the regulation of immune responses during atherosclerosis development through MHC-dependent signaling and T-cell activation [66–68]. The enrichment of junctional components - including TJP1, TJP2, and CTNND1 - further suggests coordinated modulation of cell–cell adhesion and barrier-related immune interactions [69]. Interestingly, while these junctional markers were present in the cross-sectional models, they became prominent features of the prospective model. In contrast, the prospective signature exhibited enrichment of metabolically oriented pathways, including folate biosynthesis, diabetic cataract–associated pathways, D-xylose metabolic processes, and MARVEL domain–containing protein pathways. These proteins are essential regulators of membrane apposition and tight junction organization. Their identification as prognostic markers suggests that long-term IMT progression may be associated with mechanisms related to cellular barrier organization and vesicle trafficking, rather than primarily with the acute inflammatory signaling captured by cross-sectional analyses. This shift indicates that while contemporary vascular states are characterized by overt immune activation, long-term propensity for remodeling is more closely linked to altered cellular metabolism, redox balance, and membrane structural organization.

Importantly, our temporal comparison does not support a simple transition from innate to adaptive immunity. Instead, the results are consistent with a model in which persistent myeloid immune programs interact with endothelial function, metabolic stress, and vascular aging, collectively contributing to arterial remodeling over time. In this framework, circulating immune cells likely reflect the integration of metabolic, inflammatory, and environmental signals and may serve as indicators of vascular susceptibility. Accordingly, immune transcriptional signatures detectable in peripheral blood may provide early insight into individual vascular trajectories.

From a clinical perspective, immune transcriptomic profiling holds potential implications for cardiovascular risk stratification, providing a systems-level complement to conventional clinical risk assessment. Beyond conventional static risk factors, this approach captures biological dimensions not otherwise identifiable through conventional variables, potentially helping to identify individuals predisposed to adverse vascular remodeling despite favourable baseline risk profiles. More broadly, our results support a shift from standard risk-factor evaluation toward a systems-level characterization of the immune–metabolic networks underlying cardiovascular disease development.

Several limitations should be acknowledged. The cohort size was moderate and derived from a single-center population, which may limit generalizability. Additionally, differences in ultrasound instrumentation over the 18-year span prevented a direct longitudinal comparison of absolute IMT values. Furthermore, as PBMC transcriptomics reflects composite immune populations, further studies are needed to resolve cell-type–specific transcriptional programs. Nevertheless, the exceptionally long follow-up duration, the depth of clinical phenotyping, and the integration of cross-sectional and prognostic signatures represent strengths rarely achievable in human studies of vascular remodeling.

In conclusion, this study provides robust longitudinal evidence that immune transcriptional signatures detectable in circulating blood cells are linked to subclinical vascular remodeling across nearly two decades of observation. The convergence of cross-sectional and prospective analyses indicates that arterial wall changes are associated with stable, high-level immune functional states rather than merely isolated molecular markers. Our findings support the concept that immune regulation represents an early and persistent determinant of vascular aging. Together, these findings suggest that systemic immune profiling may help refine biological cardiovascular risk stratification beyond conventional clinical factors, supporting the potential development of more personalized preventive strategies.

## Data Availability

All data produced in the present study are available upon reasonable request to the authors

## Acknowledgments

We thank Roberto Bizzotto (Institute of Neuroscience, National Research Council, Padua, Italy) and Carlo Alberto Rossi (Institute of Neuroscience, National Research Council, Padua, and Department of Cellular, Computational and Integrative Biology - CIBIO, University of Trento, Trento, Italy) for helpful comments during the development of the bioinformatic analysis pipeline used in this work.

## Appendix

**Microarray normalization**

**Figure S1.**
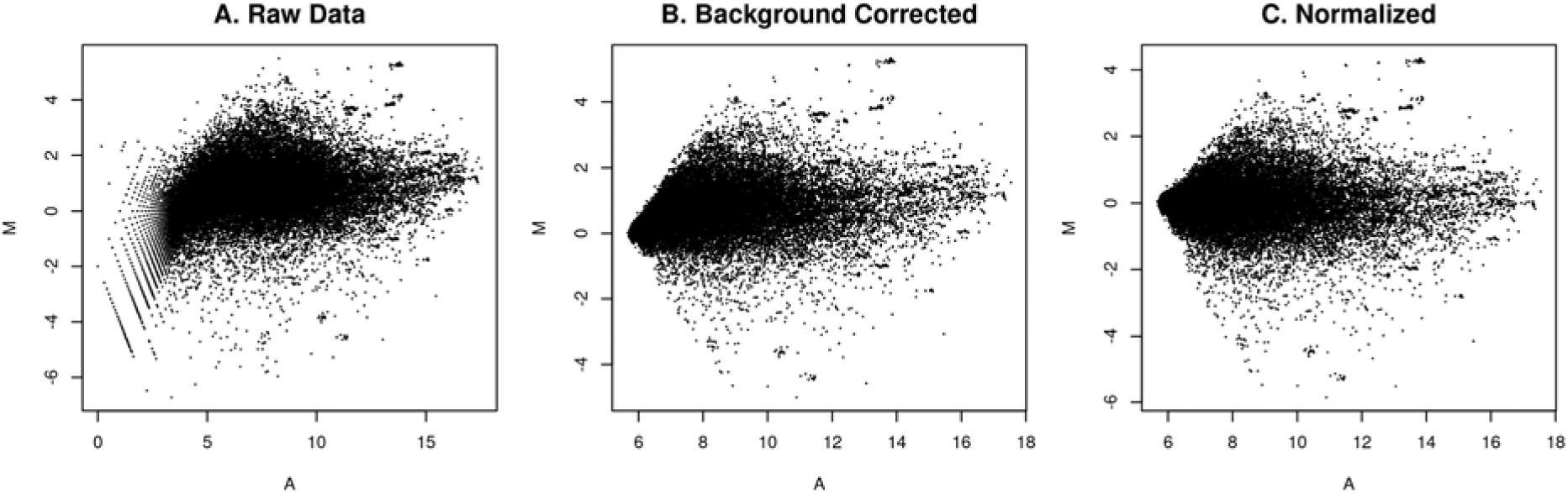
Systematic transformation of microarray expression data using MA plots. **(A)** Raw spot-level intensities exhibiting inherent background noise and non-linear dye bias. **(B)** Background-corrected data using the normexp convolution model with an offset=50. This step stabilizes the variance in the low-intensity range and prevents negative intensity values. **(C)** Data after within-array Loess normalization. The log-ratios (M) are centered around zero across the entire range of average log-intensities (A), effectively removing systematic intensity-dependent trends and preparing the data for inter-array comparison.

**Figure S2.**
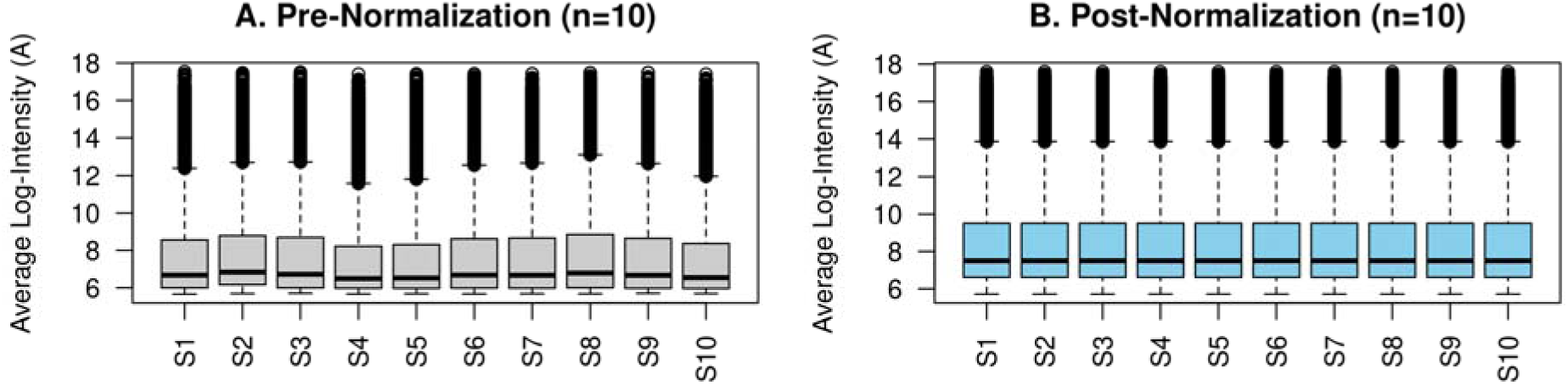
Distribution of average log-intensities (A) across representative samples. The boxplots illustrate the average log-intensity (A-values) for a subset of 10 samples to assess data quality and distributional consistency. **(A)** Pre-normalization distributions showing technical variation in intensity scales and median shifts between arrays. **(B)** Post-normalization distributions demonstrating improved alignment and stabilization of intensity ranges. The reduction in inter-array variability ensures that subsequent differential expression analysis is driven by biological signal rather than technical artifacts.

**Figure S3.**
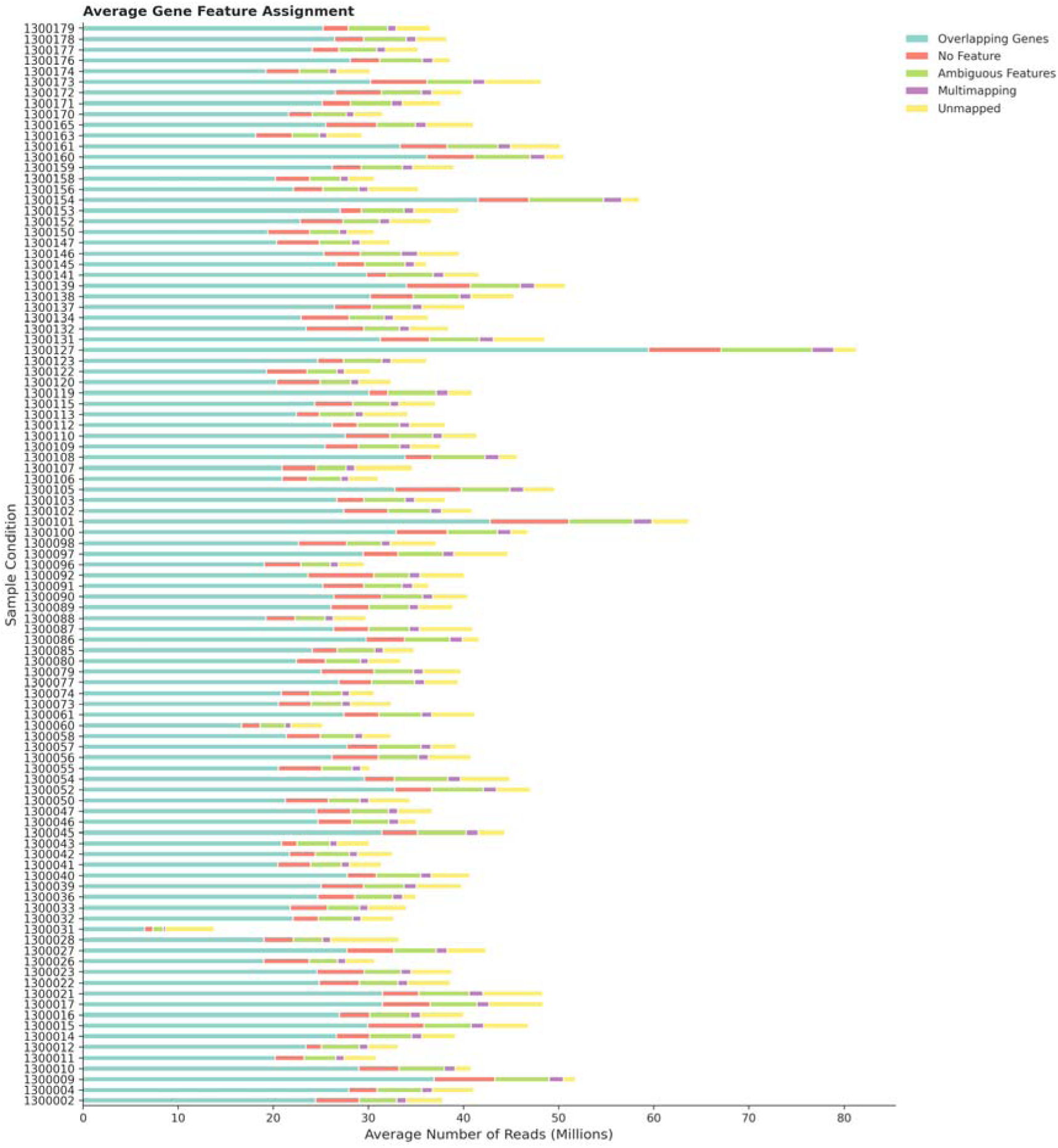
Absolute gene feature assignment summary. Stacked bars represent reads assigned to genomic features (green), or unassigned due to no feature (yellow) or ambiguous mapping (purple). Despite variations in sequencing depth, the proportional distribution of assigned reads remains uniform across the study cohort.

**Figure S4.**
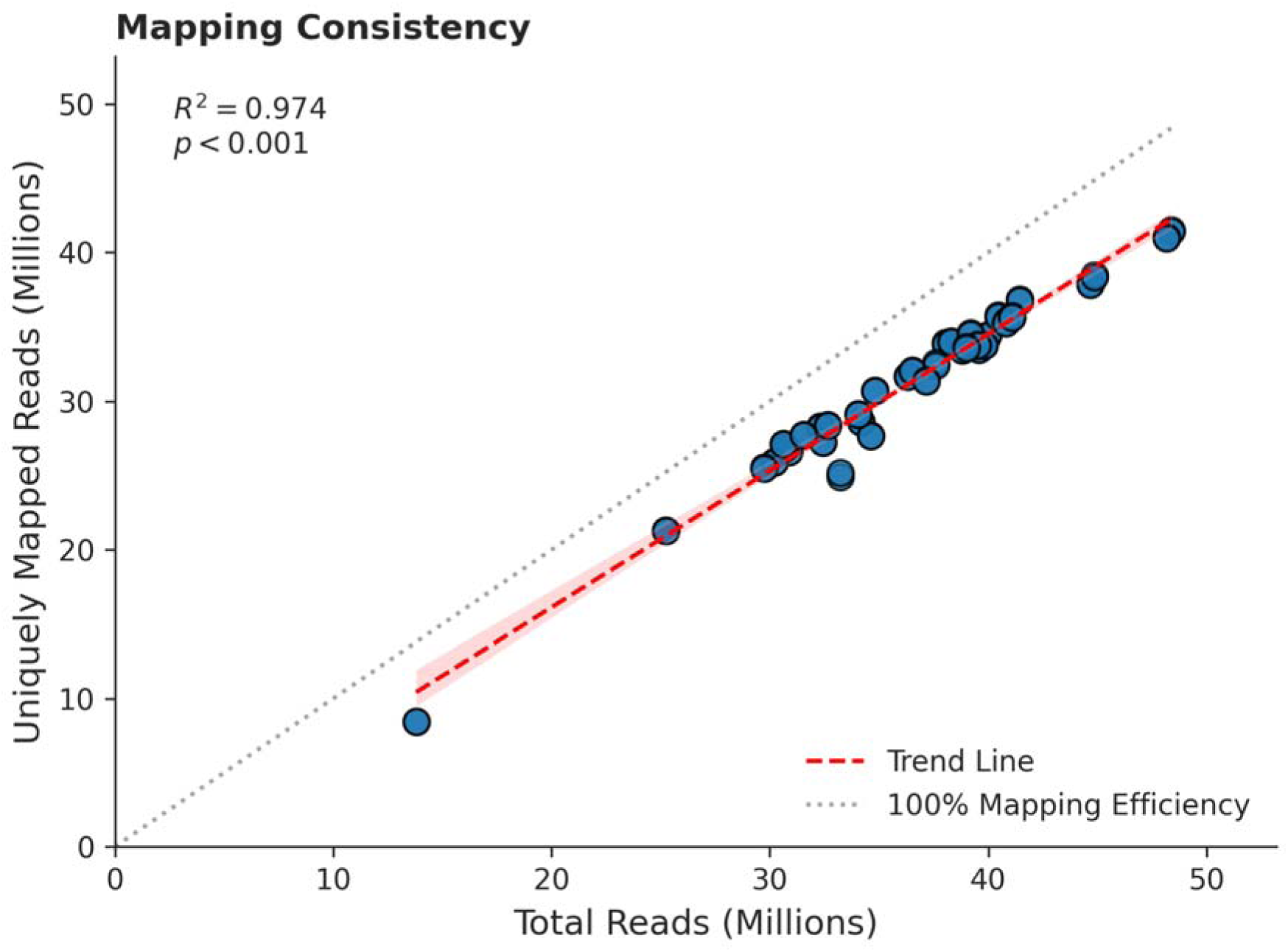
Correlation between total sequenced reads and uniquely mapped reads. The dashed red line indicates the linear regression (Trend Line), demonstrating a high coefficient of determination (R2=0.924, p<0.001). The dotted gray line represents 100% mapping efficiency. Sample **1300031** represents a technical outlier in terms of total depth (∼13.8M reads); however, it maintains a mapping efficiency consistent with the cohort trend, justifying its retention in the analysis.

**Figure S5.**
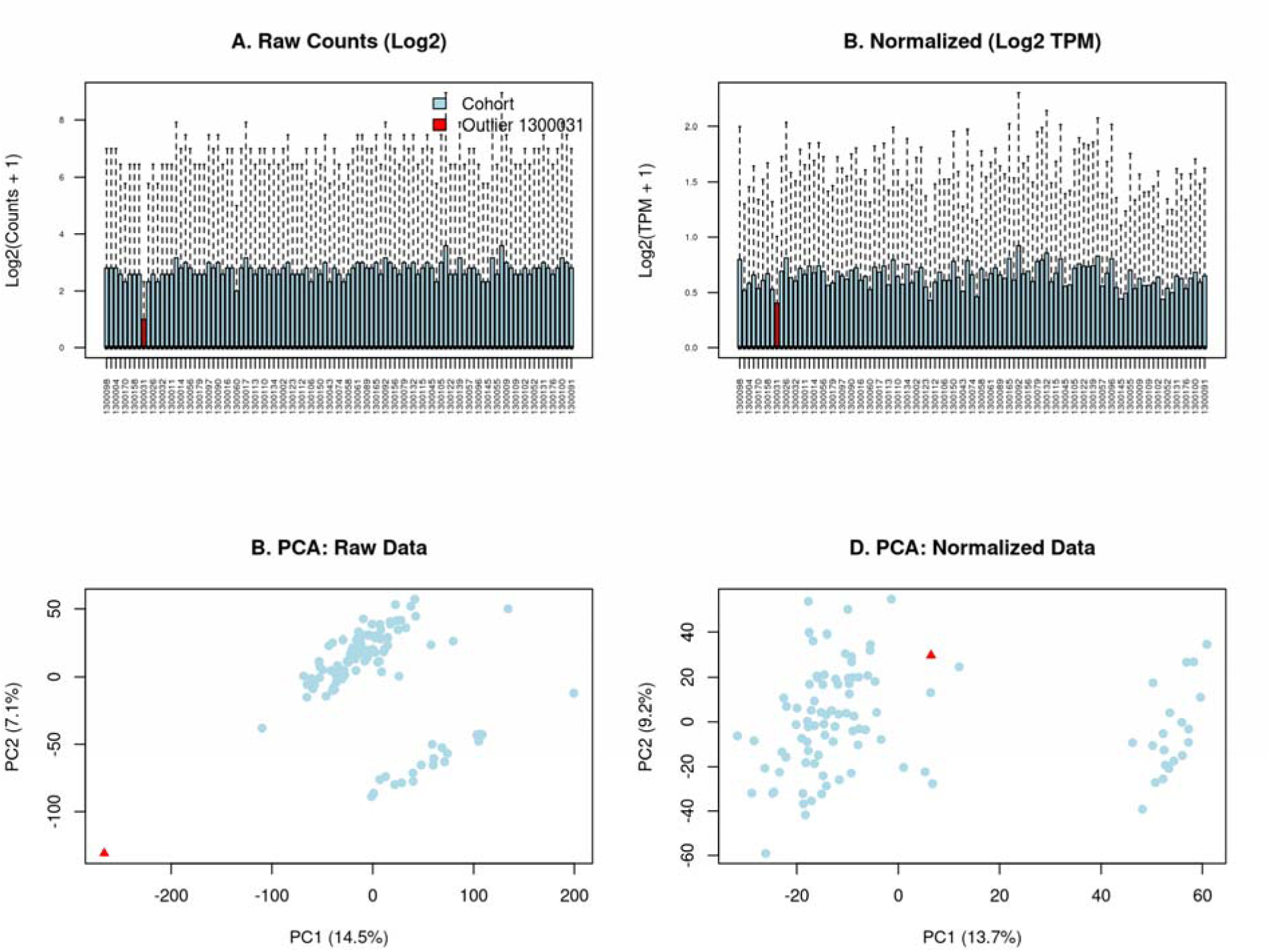
Impact of TPM normalization on cohort-wide gene expression and principal component analysis, highlighting sample 1300031. **(A)** Raw Counts (Log2) and **(B)** Normalized (Log2 TPM) bar plots of mean per-sample expression across all cohort members. Sample 1300031 (red bar) displays markedly reduced expression relative to the cohort median under raw counts, consistent with low library depth or poor-quality sequencing. Following TPM normalization, the expression level of 1300031 partially converges toward the cohort distribution, though it remains among the lower-expressing samples. **(C)** PCA of raw data reveals that sample 1300031 (red triangle) is a severe outlier, displaced prominently along both PC1 (14.5%) and PC2 (7.1%), lying far outside the main sample cluster. This separation is consistent with a global technical artifact dominating variance. (**D)** PCA of normalized data demonstrates a re-integration of sample 1300031 into the broader cohort structure. The outlier moves substantially closer to the main cluster following TPM normalization, suggesting that normalization corrects for library-size dependent technical variation.

**Figure S6.**
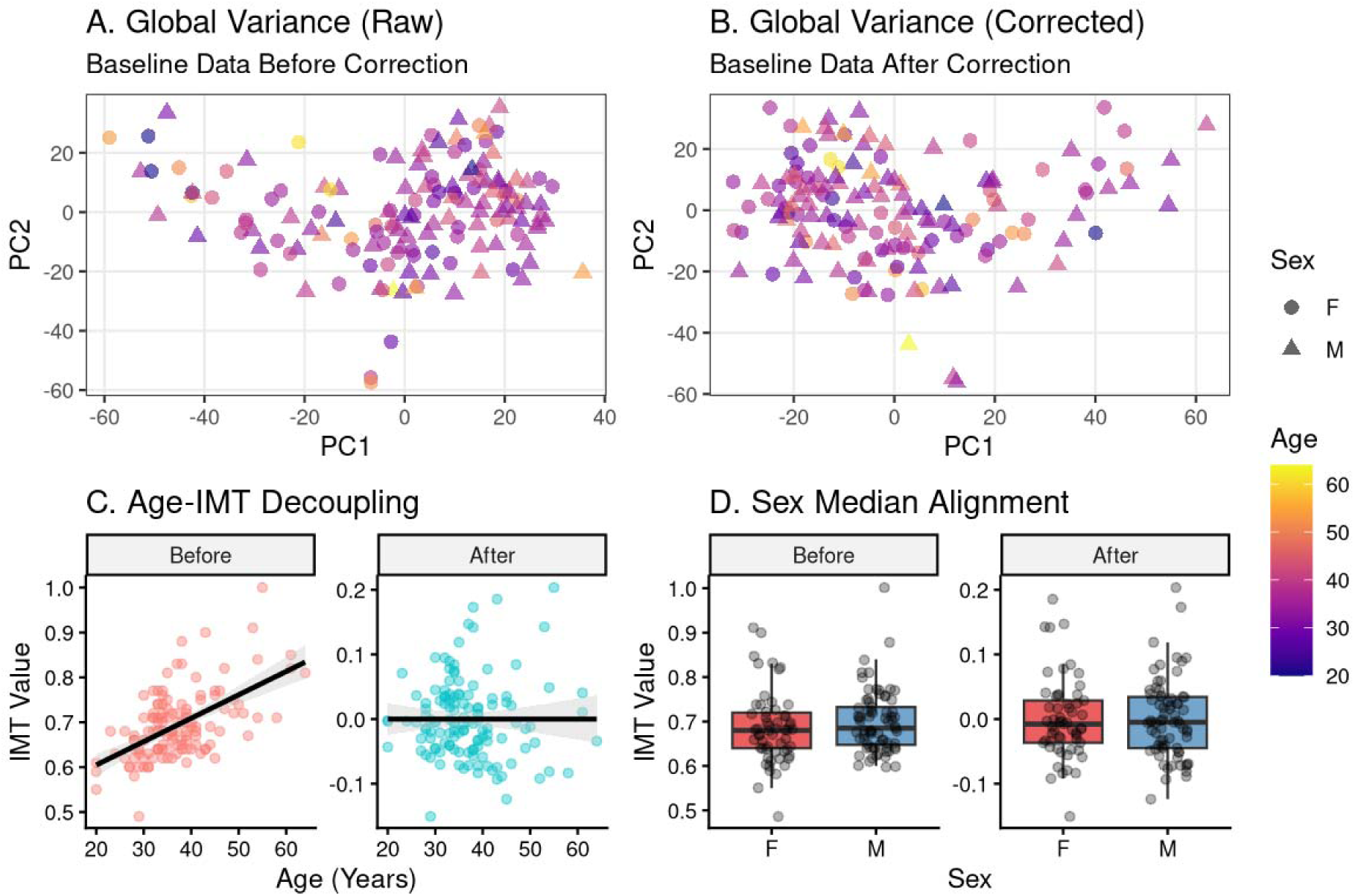
Impact of Age and Sex Covariate Correction on Gene Expression and Clinical Metadata for the Baseline Dataset. **(A–B)** Global Variance Analysis. Principal Component Analysis (PCA) of the microarray expression profiles before (A) and after (B) covariate correction. Samples are colored by Age and shaped by Sex. **(C)** Age-IMT Decoupling. Scatter plots representing the relationship between patient Age and Intima-Media Thickness (IMT). The “Before” panel shows the raw correlation inherent in the cohort; the “After” panel illustrates the reduction in the regression slope (black line) following residualization, indicating that IMT measurements have been successfully decoupled from age-related variance. (D) Sex Median Alignment. Boxplots displaying the distribution of IMT values by Sex. Post-correction (“After”), the medians and distributions are harmonized across groups, mitigating sex-based bias in the clinical metric.

**Figure S7.**
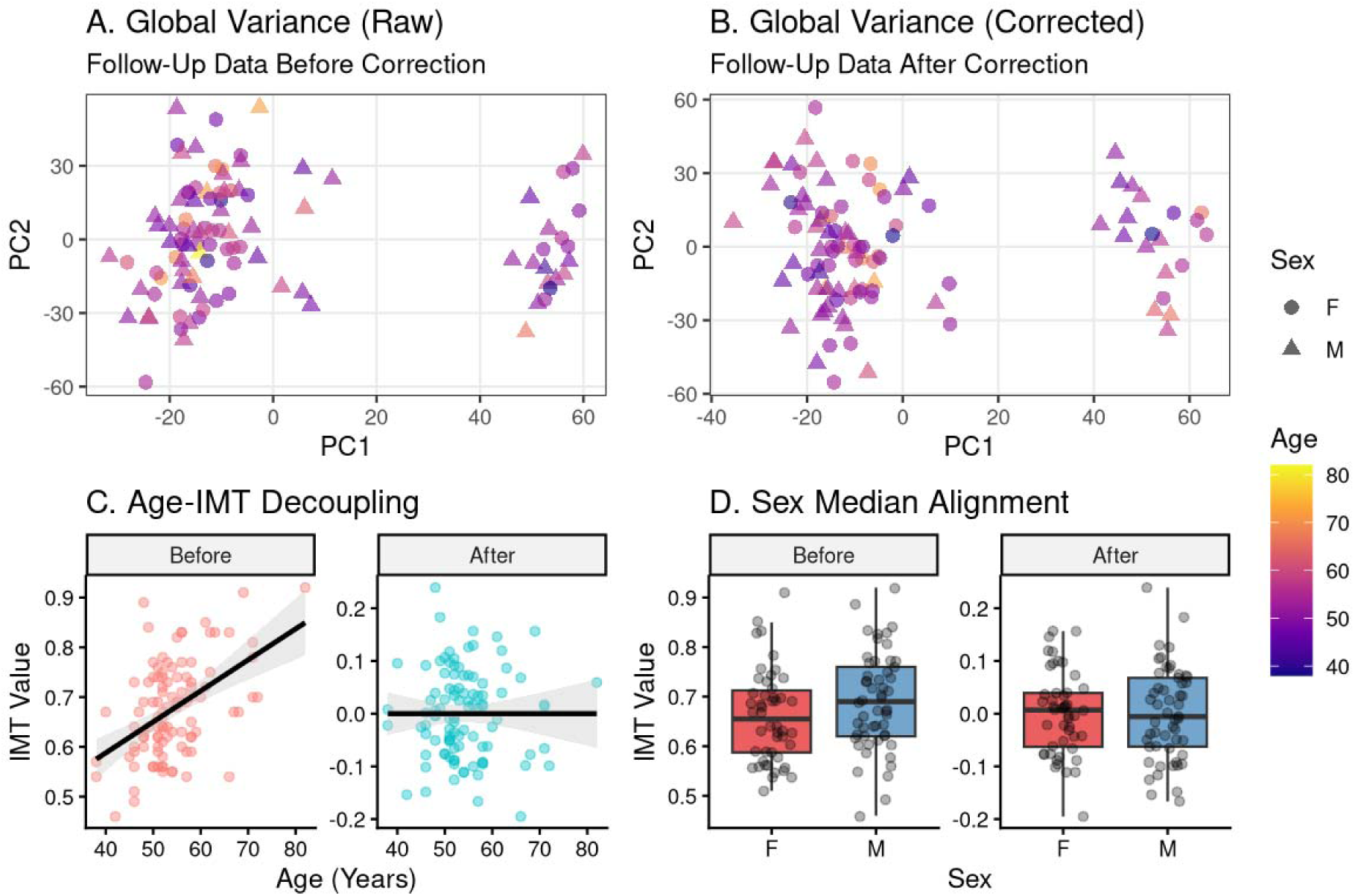
Impact of Age and Sex Covariate Correction on Gene Expression and Clinical Metadata for the Follow-Up Dataset. **(A–B)** Global Variance Analysis. Principal Component Analysis (PCA) of the rna-seq expression profiles before (A) and after (B) covariate correction. Samples are colored by Age and shaped by Sex. **(C)** Age-IMT Decoupling. Scatter plots representing the relationship between patient Age and Intima-Media Thickness (IMT). The “Before” panel shows the raw correlation inherent in the cohort; the “After” panel illustrates the reduction in the regression slope (black line) following residualization, indicating that IMT measurements have been successfully decoupled from age-related variance. (D) Sex Median Alignment. Boxplots displaying the distribution of IMT values by Sex. Post-correction (“After”), the medians and distributions are harmonized across groups, mitigating sex-based bias in the clinical metric.

